# PERFORMANCE OF αSYNUCLEIN RT-QUIC IN RELATION TO NEUROPATHOLOGICAL STAGING OF LEWY BODY DISEASE

**DOI:** 10.1101/2022.05.05.22274567

**Authors:** Sara Hall, Christina Orrù, Geidy E. Serrano, Douglas Galasko, Andrew G. Hughson, Bradley R. Groveman, Charles H. Adler, Thomas G. Beach, Byron Caughey, Oskar Hansson

## Abstract

Currently, there is a need for diagnostic markers in Lewy body disorders (LBD). αSyn RT-QuIC has emerged as a promising assay to detect misfolded α-synuclein in clinically or neuropathologically established patients with various synucleinopathies. In this study, αSyn RT-QuIC was used to analyze lumbar CSF in a clinical cohort from the Swedish BioFINDER study and postmortem ventricular CSF in a neuropathological cohort from the Arizona Study of Aging and Neurodegenerative Disorders/Brain and Body Donation Program (AZSAND/BBDP). The BioFINDER cohort included 64 PD/PDD, 15 MSA, 15 PSP, 47 controls and two controls who later converted to PD/DLB. The neuropathological cohort included 101 cases with different brain disorders, including LBD and controls. In the BioFINDER cohort αSyn RT-QuIC identified LBD (i.e. PD, PDD and converters) vs. controls with a sensitivity of 95% and a specificity of 83%. The two controls that converted to LBD were αSyn RT-QuIC positive. Within the AZSAND/BBDP cohort, αSyn RT-QuIC identified neuropathologically verified “standard LBD” (i.e. PD, PD with AD and DLB; n=25) vs. no LB pathology (n=53) with high sensitivity (100%) and specificity (94%). Only 57% were αSyn RT-QuIC positive in the subgroup with “non-standard” LBD (i.e., AD with Lewy Bodies not meeting criteria for DLB or PD, and incidental LBD, n=23). Furthermore, αSyn RT-QuIC reliably identified cases with LB pathology in the cortex (97% sensitivity) vs. cases with no LBs or LBs present only in the olfactory bulb (93% specificity). However, the sensitivity was low, only 50%, for cases with LB pathology restricted to the brainstem or amygdala, not affecting the allocortex or neocortex. In conclusion, αSyn RT-QuIC of CSF samples is highly sensitive and specific for identifying cases with clinicopathologically-defined Lewy body disorders and shows a lower sensitivity for non-standard LBD or asymptomatic LBD or in cases with modest LB pathology not affecting the cortex.

## Introduction

The clinical diagnosis of Parkinson’s disease (PD) can be difficult due to its heterogenous presentation and clinical overlap with atypical parkinsonian disorders, especially early in the disease course. Indeed, the diagnostic accuracy for PD has been found to be as low as 73.8% (67.8-79.6) by non-experts but only slightly better by movement disorders specialist with an accuracy of 79.6% (46-95.1) at the initial assessment, particularly when disease duration is less than 5 years [2, 3, 7, 52]. The differential diagnosis of the atypical parkinsonian disorders, multiple system atrophy (MSA), progressive nuclear palsy (PSP), and corticobasal degeneration (CBD) are even more difficult, with generally acceptable specificity but low sensitivity[33, 40].

There are to date no disease-modifying therapies available in PD. New disease-modifying therapies are likely to be most efficient early on in the disease process, before neuronal damage is irreversible [58]. The lack of clear and reliable biomarkers that can identify individuals with PD has been considered a great barrier to the development of disease-modifying treatments [17]. There is thus an urgent need for early and accurate biomarkers for Parkinson’s disease.

Although results consistently have shown decreased levels of CSF unmodified α-synuclein (α-syn) in PD, but also PD with dementia (PDD), dementia with Lewy bodies (DLB), and MSA compared with controls, the reduction is modest, with a broad overlap with controls, and subsequently has failed to adequately discriminate between PD and controls [29, 45, 47], hampering its usefulness in clinical trials and practice.

Ultrasensitive seed amplification assays are methods originally developed for the detection of misfolded prion proteins and prion-like proteins. Over the last several years αSyn RT-QuIC and related α-syn seed amplification assays have emerged as possible methods for detecting misfolded forms of synuclein in CSF, exploiting the prion-like propagation mechanism of pathological α-syn aggregates. Previous studies have shown a high sensitivity of > 92% for clinicopathologically verified Lewy body disorders (LBD), i.e. PD, PDD and DLB, compared to controls/non-synucleinopathies, and a specificity of > 95% [5, 14, 23, 55]. The diagnostic accuracy has predictably been somewhat lower in clinical cohorts with sensitivity > 93% and specificity > 82% [14, 28, 39, 46, 56]. Among αSyn RT-QuIC assays, there are those with particularly short overall assay times of ∼1-2 days, without compromising diagnostic performance [16, 28, 46, 51, 54-56, 64].

In this study we use a rapid αSyn RT-QuIC assay to test CSF samples from participants with clinically diagnosed PD, PDD, MSA, PSP and controls in the longitudinal Swedish BioFINDER study[30] and participants from a well characterized cohort of neuropathologically-verified cases with different brain disorders including cases and controls from the Arizona Study of Aging and Neurodegenerative Disorders/Brain and Body Donation Program (AZSAND/BBDP)[9]. Previous neuropathological studies have mainly compared “ideal” groups of cases, i.e. controls with no LB disease versus neuropathologically verified and clinically-manifest (“standard LBD”: PD, PD with Alzheimer’s disease, PDD and DLB). In this study, we investigated not only individuals with standard LBD and controls, but also those with non-standard LBD (i.e., the cases with Lewy bodies at autopsy but not meeting clinicopathological consensus criteria for DLB or PD), including cases with Alzheimer’s disease with Lewy bodies not meeting criteria for DLB or PD and cases with incidental Lewy body disease (ILBD). Additionally, we investigated CSF αSyn RT-QuIC results in relation to i) the LB stage, ii) the LB density and iii) the LB distribution in ten selected brain regions irrespective of the clinical and neuropathological diagnosis.

## Methods

### Participants in the BioFINDER cohort

The study was performed at the Clinic of Neurology, Skåne University hospital, Sweden as part of the Swedish BioFINDER Study (www.biofinder.se) [30]. In this convenience study, the study participants are primarily recruited from the southern region of Sweden and recruited between 2008 and 2017. Patients with PD (n=50) met the NINDS Diagnostic Criteria for PD [25]. Patients with PDD (n=14) also met criteria for PDD at baseline [21]. Patients with MSA (n=15) met the consensus statement by Gilman et al. [26]. Patients with PSP (n=15) met the criteria according to the report of the National Institute of Neurological Disorders and Stroke–Society for Progressive Supranuclear Palsy International Workshop [41]. All controls (n=47) underwent cognitive testing and neurologic examination by a medical doctor and individuals with objective cognitive or parkinsonian symptoms were not included. Participants are followed with repeated assessment for up to 10 years. Two individuals initially included as controls converted to clinical LBD during follow-up. One was diagnosed with PD after 5.5 years follow-up and one was diagnosed with DLB after 3.5 years follow-up. Exclusion criteria were i) age above 85 years, ii) presence of generalized malignancy, iii) ongoing or earlier advanced abuse of alcohol or illicit drugs, iv) presence of clinically-diagnosed Alzheimer’s dementia, vascular dementia, frontotemporal lobe dementia, v) presence of severe psychiatric disorders, vi) presence of other severe neurological disease, vii) participation in clinical drug trial within the last 30 days.

All participants gave written informed consent before entering the study. The study procedure was approved by the local ethics committee at Lund University Sweden and conducted according to the Helsinki Declaration.

A thorough medical history was taken and the participants underwent extensive testing. Participants were examined by a physician experienced in movement disorders and a registered research nurse using, among other scales, the Unified Parkinson’s Disease Rating Scale (UPDRS) -3, the Hoehn & Yahr scale and the Mini Mental State Examination (MMSE) [22, 24, 32].

CSF samples were obtained by lumbar puncture in the L3/L4 or L4/L5 interspace with patient non-fasting. The samples were collected in polypropylene tubes and gently mixed to avoid gradient effects.

All samples were centrifuged within 30 minutes at +4°C at 2000g for 10 min to remove cells and debris, and then stored in aliquots at −80°C pending biochemical analysis. The procedure followed the Alzheimer’s Association Flow Chart for CSF biomarkers [13].

### Participants in AZSAND/BBDP cohort

The neuropathology cohort consisted of neuropathologically classified participants (n=101) from the Arizona Study of Aging and Neurodegenerative Disorders (AZSAND), an antemortem-postmortem donor cohort with dates of enrollment from 2007 to 2019. Autopsies were performed by the Banner Sun Health Research Institute Brain and Body Donation program (BBDP) [9, 11].

Neuropathological diagnosis of PD was based on a combination of established neuropathologic criteria [8, 11, 20] and a clinical diagnosis of parkinsonism. DLB diagnosis was defined as a clinical diagnosis of dementia with an intermediate or high likelihood of DLB by the third meeting of the Dementia with Lewy Bodies Consortium [43]

Cases were classified using the Unified Staging System for Lewy Body Disorders (USSLBD) i.e., cases with LBs present were classified into LB stages: I. Olfactory Bulb Only; IIa Brainstem Predominant; IIb Limbic Predominant; III Brainstem and Limbic; IV Neocortical [8]. Lewy Body (LB) density score was assessed in all cases. LB density score is a semi quantitative score of 0-4 in 10 different brain regions (olfactory bulb and tract, medulla at the level of the 9^th^ and 10^th^ cranial nerve nuclei, pons at the level of the locus ceruleus, amygdala, substantia nigra, transentorhinal area, cingulate gyrus at level just posterior to genu of corpus callosum, middle temporal gyrus, middle frontal gyrus, inferior parietal lobule) yielding a maximum score of 40. Neuronal perikaryal cytoplasmic staining, neurites and puncta were considered together, using the templates provided by the Dementia with Lewy Bodies Consortium [43]. The immunohistochemical method used an antibody against phosphorylated synuclein, as previously described[8].

Cases were classified as having PD or DLB, or ILBD (incidental Lewy body disease) in the case of controls without parkinsonism or dementia, when Lewy bodies were present on neuropathological examination but cases did not meet clinicopathological diagnostic criteria for either PD or DLB.

PSP, CBD and MSA were diagnosed according to previously published criteria [18, 19, 27, 31]. Neuropathological diagnosis of AD was based on National Institute on Aging–Reagan Institute (NIA-RI) criteria [1] which are dependent on the Consortium to Establish a Registry for Alzheimer disease (CERAD) neuritic plaque [44] and Braak (neurofibrillary tau-tangle) stage [15]. National Institute on Aging-Alzheimer’s Association criteria[34] were not used due to many cases who had autopsy prior to these newer criteria. The major difference between the two sets of criteria are the addition of Thal amyloid phase to the newer criteria but this may not improve clinicopathological correlations[59]. Controls were cases without dementia or parkinsonism during life and without a major neuropathological diagnosis.

Histopathological scoring was performed blinded to clinical and neuropathological diagnosis. Amyloid plaque and neurofibrillary tangle density were both graded and staged at standard sites in frontal, temporal and parietal lobes and hippocampal CA1 region and entorhinal region using a semi-quantitative score of 0-3 based on the CERAD templates, yielding a total score of 15 [9, 44]. Immunohistochemical staining for phosphorylated TDP-43 was performed in 38 of the 101 cases as previously described [4, 9].

Post mortm CSF was drawn from the lateral ventricles prior to removing the brain. The CSF was then ejected into 15 mL disposable polyethylene tubes. CSF was centrifuged at 2,000 g for 10 minutes at 24 C and supernatants were aliquoted into 0.5 mL polyethylene microcentrifuge tubes and stored frozen at −80°C [9].

All participants had signed written informed consent. Ethical approval was given by Banner Health-designated Institutional Review Boards; currently the Western Institutional Review Board of Puyallup, Washington.

### αSyn RT-QuIC analyses

αSyn RT-QuIC analyses were performed blinded to clinical and neuropathological status and diagnosis of the patient. The preparation of the K23Q recombinant α-synuclein was done as previously described [57]. The αSyn RT-QuIC analyses was done as previously described [46]. Briefly, reactions were performed in black 96-well plates with a clear bottom (Nalgene Nunc International). Each well was preloaded with six glass beads (0.8 mm in diameter, OPS Diagnostics). Quadruplicate reactions were seeded with 15 μL of CSF. Prior to the addition of CSF, each RT-QuIC reaction mix was 85 μL of solution [28] with final reaction concentrations of 40 mM sodium phosphate buffer, 170 mM NaCl, 0.1 mg/mL K23Q recombinant αSyn (filtered through a 100 kD MWCO filter immediately prior to use), 10 μM thioflavin T (ThT) and 0.0015% SDS. The plates were closed with a plate sealer film (Nalgene Nunc International) and incubated at 42°C in a BMG FLUOstar Omega plate reader. The plates were then incubated for at least 48 h and subjected to cycles of 1 min shaking (400 rpm double orbital) and 1 min rest for at least 48 h. ThT fluorescence measurements (450 +/− 10 nm excitation and 480 +/− 10 nm emission; bottom read) were taken every 45 min with fluorimeter gain settings adjusted to maintain fluorescence responses within an unsaturated range (in most cases). The fluorescence threshold was calculated individually for each 96-well plate to account for differences between plate readers. Positive reactions were those exceeding 10% of the maximum value obtained on the same plate (e.g from positive controls). For a sample to be considered positive overall, the fluorescent signal had to exceed the threshold in at least 50% of the replicate wells (e.g. ≥2 of 4) within 48 h.

### Statistical analyses

Mann-Whitney U test was used for comparison of continuous and ordinal variables between groups and chi-square test for dichotomous variables. p<0.05 (two-sided) was considered statistically significant. Sensitivity and specificity of the αSyn RT-QuIC results were calculated for diagnoses in the clinical BioFINDER cohort and between the presence / absence of any LB pathology, LB stage, LB density score, LB distribution and clinicopathological diagnosis in the AZSAND/BBDP cohort. Univariate associations between two continuous or ordinal variables were analyzed using Spearman ρ. SPSS (version 27; SPSS Inc., Chicago, Illinois) was used for statistical analyses and figures.

## Results

### Clinical BioFINDER cohort

There were no sex differences between the diagnostic groups and no significant difference between men and women in αSyn RT-QuIC results (Table 1). There was no significant difference in age between αSyn RT-QuIC positive and negative individuals.

**Table 1.**
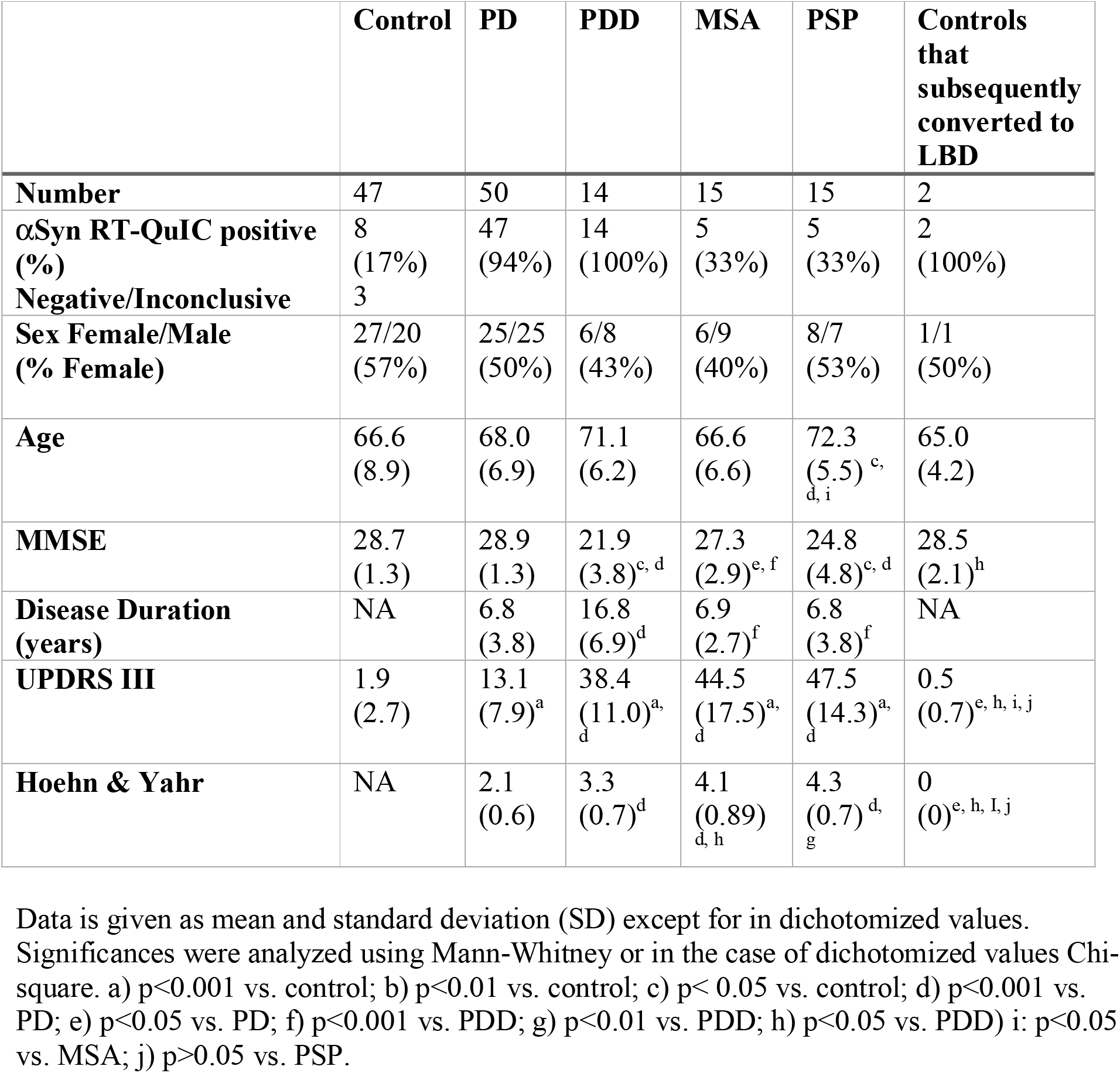
Patient characteristics, BioFINDER cohort.

|  | **Control** | **PD** | **PDD** | **MSA** | **PSP** | **Controls that subsequently converted to LBD** |
| --- | --- | --- | --- | --- | --- | --- |
| **Number** | 47 | 50 | 14 | 15 | 15 | 2 |
| **αSyn RT-QuIC positive (%)** | 8 (17%) | 47 (94%) | 14 (100%) | 5 (33%) | 5 (33%) | 2 (100%) |
| **Negative/Inconclusive** | 3 |  |  |  |  |  |
| **Sex Female/Male (% Female)** | 27/20 (57%) | 25/25 (50%) | 6/8 (43%) | 6/9 (40%) | 8/7 (53%) | 1/1 (50%) |
| **Age** | 66.6 (8.9) | 68.0 (6.9) | 71.1 (6.2) | 66.6 (6.6) | 72.3 (5.5)<sup>c, d, i</sup> | 65.0 (4.2) |
| **MMSE** | 28.7 (1.3) | 28.9 (1.3) | 21.9 (3.8)<sup>c, d</sup> | 27.3 (2.9)<sup>e, f</sup> | 24.8 (4.8)<sup>c, d</sup> | 28.5 (2.1)<sup>h</sup> |
| **Disease Duration (years)** | NA | 6.8 (3.8) | 16.8 (6.9)<sup>d</sup> | 6.9 (2.7)<sup>f</sup> | 6.8 (3.8)<sup>f</sup> | NA |
| **UPDRS III** | 1.9 (2.7) | 13.1 (7.9)<sup>a</sup> | 38.4 (11.0)<sup>a, d</sup> | 44.5 (17.5)<sup>a, d</sup> | 47.5 (14.3)<sup>a, d</sup> | 0.5 (0.7)<sup>e, h, i, j</sup> |
| **Hoehn & Yahr** | NA | 2.1 (0.6) | 3.3 (0.7)<sup>d</sup> | 4.1 (0.89)<sup>d, h</sup> | 4.3 (0.7)<sup>d, g</sup> | 0 (0)<sup>e, h, I, j</sup> |

Data is given as mean and standard deviation (SD) except for in dichotomized values. Significances were analyzed using Mann-Whitney or in the case of dichotomized values Chi-square. a) p<0.001 vs. control; b) p<0.01 vs. control; c) p< 0.05 vs. control; d) p<0.001 vs. PD; e) p<0.05 vs. PD; f) p<0.001 vs. PDD; g) p<0.01 vs. PDD; h) p<0.05 vs. PDD) i: p<0.05 vs. MSA; j) p>0.05 vs. PSP.

#### CSF αSyn RT-QuIC vs clinical diagnosis

In the clinically diagnosed BioFINDER study, 94% of PD patients and 100% of PDD patients were αSyn RT-QuIC positive compared with 17% of controls (n=8) and 33% of PSP patients. Further, 33% of those with clinically diagnosed MSA were αSyn RT-QuIC positive.

Interestingly, both individuals who were included as controls but subsequently developed clinically diagnosed LBD (PD and DLB respectively) during follow up had positive αSyn RT-QuIC result.

In this clinical cohort, αSyn RT-QuIC could discriminate between controls and non-demented PD with a sensitivity of 94% and specificity of 83%. When including PD, PDD and individuals who later converted to PD, the sensitivity was slightly higher (95%). However, the three αSyn RT-QuIC negative had disease duration of ≤5 years yielding a sensitity for cases with early PD (≤5 years) including converters of 90% and 100% in cases with advanced PD, including PDD (>5) years. Furthermore, αSyn RT-QuIC discriminated between LBD vs. MSA and PSP with a sensitivity of 95% and a specificity of 67%.

### AZSAND/BBDP cohort

There was no significant difference between men and women regarding CSF αSyn RT-QuIC results in this neuropathology-based cohort (Table 2). As expected, there was a strong correlation between LB stage and LB density total score (Rs=0.990, p<0.001).

**Table 2.** Patient characteristics, AZSAND/BBDP cohort.

|  | **αSyn RT-QuIC**<br>**positive** | **αSyn RT-QuIC**<br>**negative** | **p-value** |
| --- | --- | --- | --- |
| **Number** | 41 | 60 |  |
| **Age at death, Mean (SD)** | 83 (7.0) | 86 (8.6) | **0.002** |
| **Sex Male/Female** | 31/10 | 35/25 | 0.073 |
| **Lewy Body positive** | 38 (93%) | 10 (17%) | **<0.001** |
| **Lewy Body density total score (max 40)**<br>**Mean (SD)** | 21 (11)<br>1 missing | 1.05 (2.9) | **<0.001** |
| **Lewy Body stage** |  |  |  |
| • **0** | 3 | 50 |  |
| • **I (olfactory bulb only)** | 1 | 2 |  |
| • **IIa (brainstem predominant)** | 2 | 3 |  |
| • **IIb (limbic predominant)** | 7 | 4 |  |
| • **III (brainstem/limbic)** | 16 | 1 |  |
| • **IV (neocortical)** | 12 | 0 |  |
| **Total senile plaque density score (0-15)**<br>**Mean (SD)** | 8.05 (6.19) | 7.16 (6.25) | p=0.427 |
| **Total neurofibrillary tangle density**<br>**score (0-15), Mean (SD)** | 7.70 (3.73) | 6.95 (2.94) | p=0.703 |
| **TDP-43 pathology Yes/No (missing)** | 3/10 (28) | 7/18 (35) | p=0.744 |

Significances are calculated using Mann-Whitney (continuous and ordinal variables) or $\chi^2$ (dichotomized variables).

#### CSF αSyn RT-QuIC status by LB stage, AZSAND/BBDP cohort

The frequency of αSyn RT-QuIC positive cases in each LB stage are given in Fig. 1. Of the αSyn RT-QuIC positive cases, 93% (38 out of 41) were LB positive (LB stage I-IV) compared to 17% (10 out of 60) cases in the αSyn RT-QuIC negative group (Table 2). Virtually all of those in LB stage III-IV were αSyn RT-QuIC positive (28 out of 29) and in LB stage 0-1 most were negative (52 out of 56); yielding a sensitivity of 97% and specificity of 93% when comparing LB stage III-IV vs LB stage 0-1 (Table 2; Fig. 1). 2/3 with olfactory bulb only (LB stage I) were αSyn RT-QuIC negative and including LB stage 0 only, the specificity was 94%. However, the sensitivity for detecting LB pathology in stage IIa-IIb was only 56% (9 out of 16). Of note is that the single LB stage III case that was αSyn RT-QuIC negative had a relatively low LB density score of 9 compared to a mean of 23.5 (SD 5.1) in all cases with LB stage III. Of the 16 with LB stage IIa-IIb none had clinical PD or DLB diagnosis. None of the 9 αSyn RT-QuIC positive cases had a parkinsonian clinical diagnosis and although the 16 cases with LB stage IIa-IIb had a mean UPDRS motor score of 16.4 (SD 21.9) there was no significant difference between αSyn RT-QuIC positive and negative cases.

**Fig. 1.**
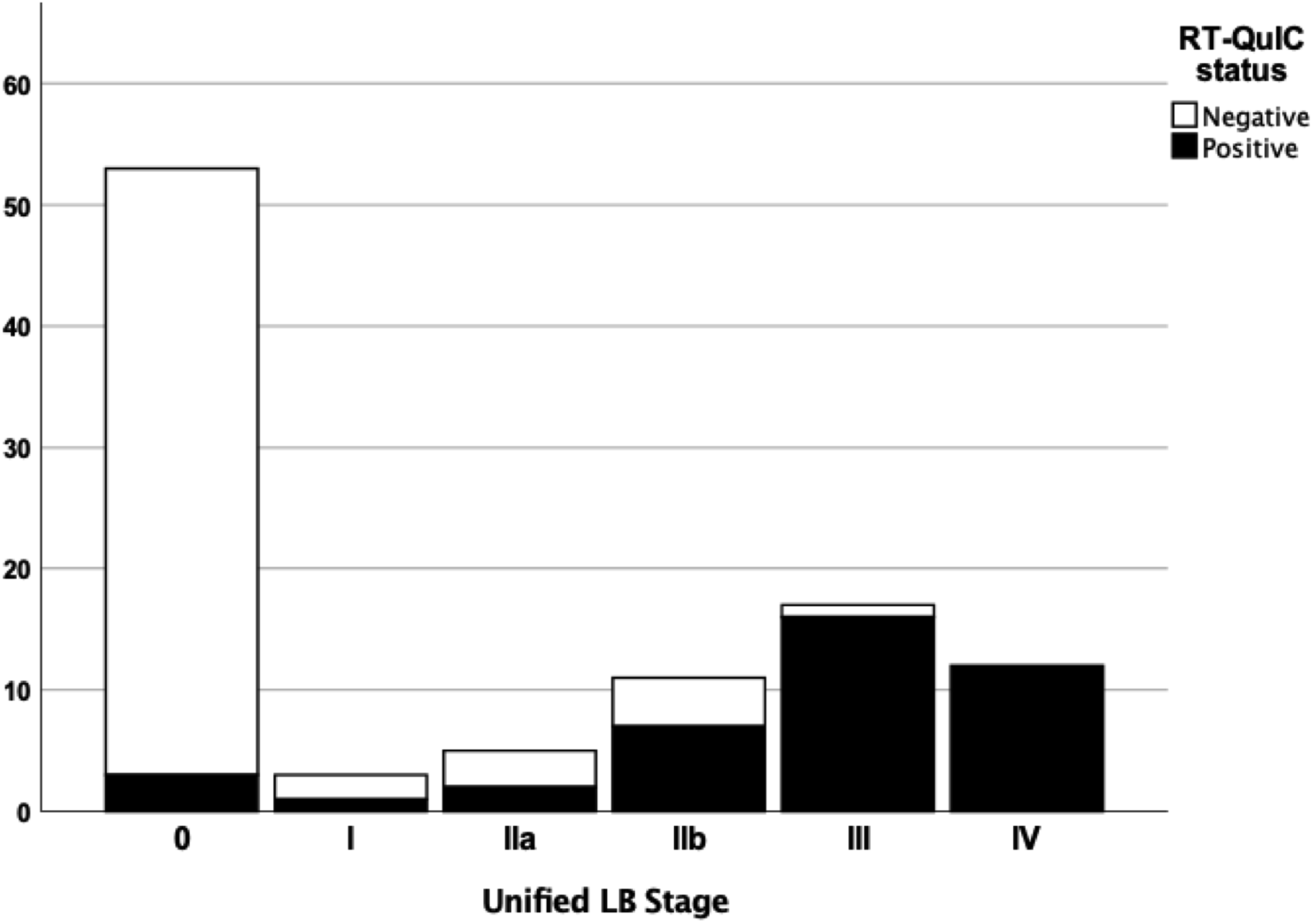
Stacked bar chart depicting αSyn RT-QuIC status by LB stage in the neuropathology-based AZSAND/BBDP cohort. 97% of the cases with LB stage III-IV were αSyn RT-QuIC positive and 93% of the cases in LB stage 0-1 were αSyn RT-QuIC negative. However, only 56% of cases with LB pathology in stage IIa-IIb were αSyn RT-QuIC positive

#### CSF αSyn RT-QuIC status by LB distribution, AZSAND/BBDP cohort

When comparing the CSF αSyn RT-QuIC results more carefully to the distribution of LB pathology in the brain we found that 97% of cases with LB pathology in the cortex (allocortex and/or neocortex) were αSyn RT-QuIC positive (30 out of 31; Fig. 2). The one αSyn RT-QuIC-negative case with cortical LBs was at the LB stage IIb and had a LB density score of only 1 in the cingulum (due to one single LB) and no other cortical involvement. Of the 14 cases with LB pathology in the brainstem and/or amygdala, but with no cortical involvement, 50% were αSyn RT-QuIC positive. (Fig. 2). We next subdivided these 14 cases further into brainstem only (n=2), amygdala only (n=7) or those with LB pathology restricted to both brain stem and amygdala (n=5). We found that 2 out of 2 “brainstem only” were negative, 2 out of 7 “amygdala only” were negative, and 3 out of 5 with “LB pathology restricted to both amygdala and brainstem” were negative.

**Fig. 2.**
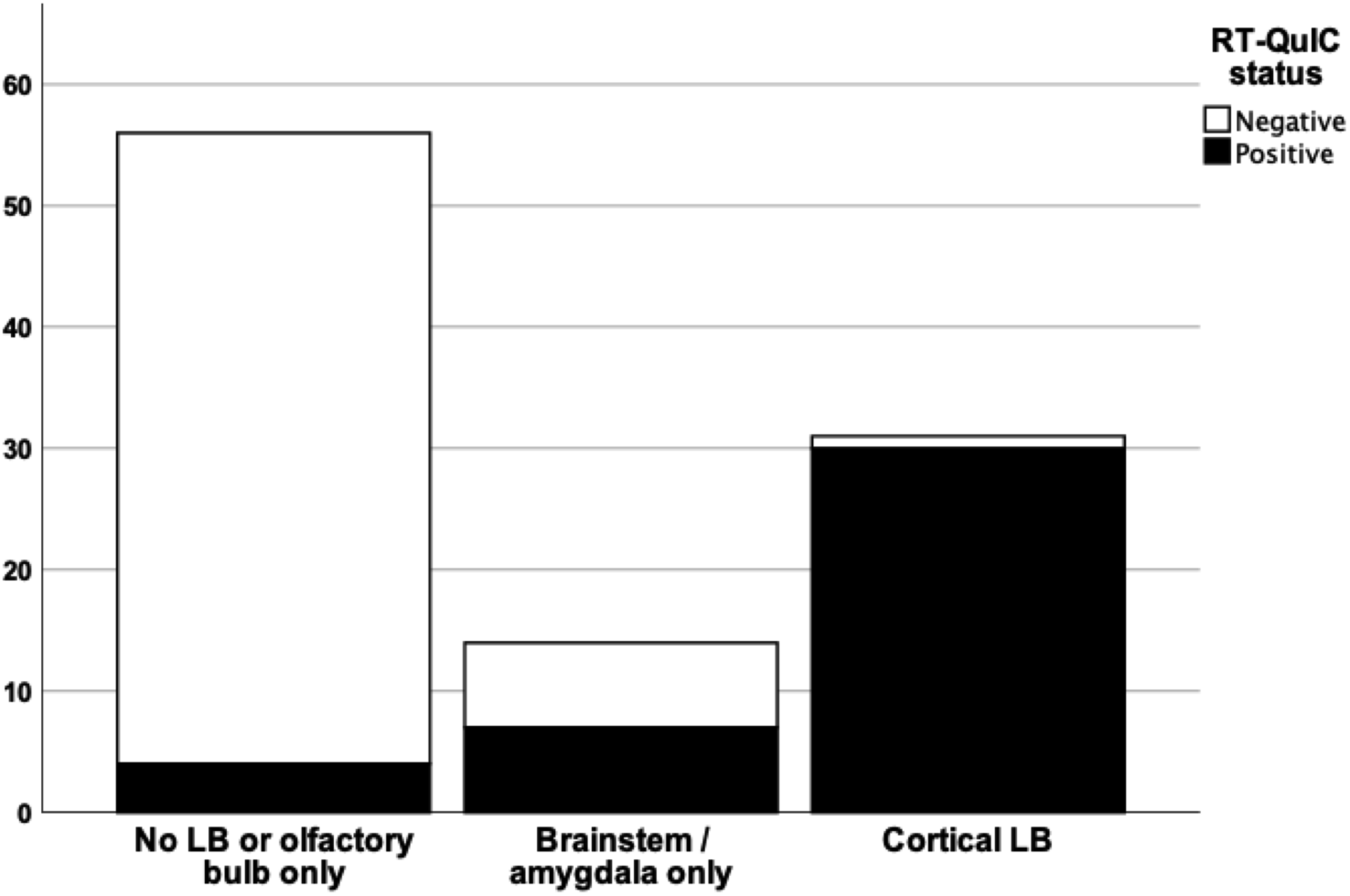
Stacked bar chart depicting αSyn RT-QuIC status by LB distribution in the neuropathology-based AZSAND/BBDP cohort. 97% of cases with LB pathology in the cortex (allocortex and/or neocortex) were αSyn RT-QuIC positive whereas 93% of cases with no LBs or LB restricted to the olfactory bulb only were αSyn RT-QuIC negative. Of the cases with LB pathology in the brainstem and/or amygdala, but with no cortical involvement, 50% were αSyn RT-QuIC positive

#### CSF αSyn RT-QuIC status by LB density, AZSAND/BBDP cohort

When using the total LB density score (established by summing the regional density scores from the ten predefined regions), the αSyn RT-QuIC negative cases had significantly lower LB density compared with αSyn RT-QuIC positive cases (p<0.001) (Table 2). αSyn RT-QuIC identified cases with total LB density score >10 with a sensitivity of 97% (29 out of 30 were positive) and cases with total LB density score 0-4 with a specificity of 93% (55 out of 59 were negative). Of the cases with an intermediate LB density score of 5-10, however, only 64% (7 out of 11) were αSyn RT-QuIC positive (Fig. 3).

**Fig. 3.**
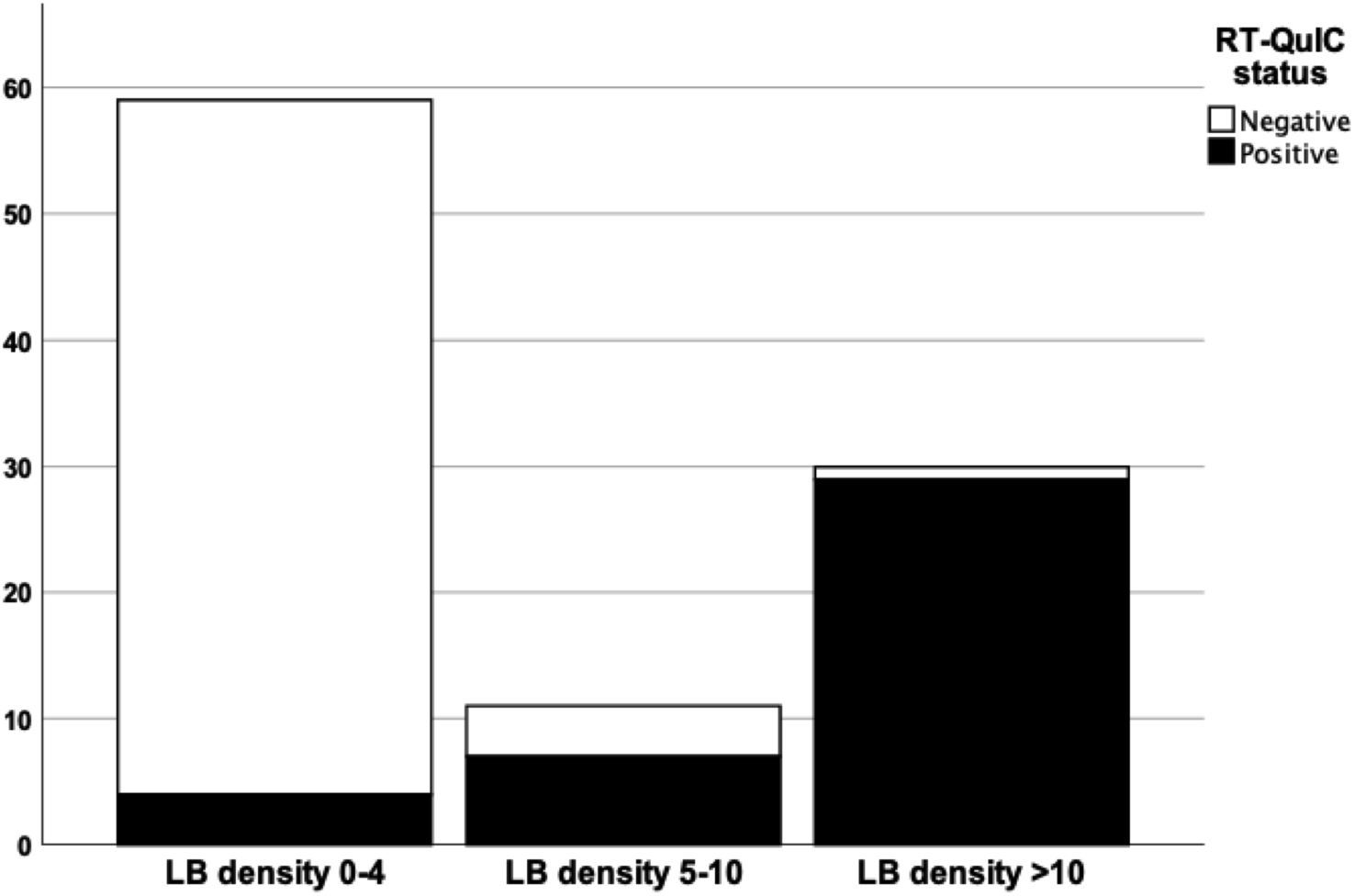
Stacked bar chart depicting αSyn RT-QuIC status by LB density in the neuropathology-based AZSAND/BBDP cohort. 97% of cases with total LB density score >10 score (established by summing the regional density scores from the ten predefined regions yielding a maximum score of 40) were αSyn RT-QuIC positive. 93% of cases with total LB density score of 0-4 were αSyn RT-QuIC negative. Only 64% of cases with an intermediate LB density score of 5-10 were αSyn RT-QuIC positive

#### CSF αSyn RT-QuIC by clinicopathological diagnosis, AZSAND/BBDP cohort

CSF αSyn RT-QuIC identified neuropathologically verified “standard LBD” (i.e. PD, PD with AD and DLB with AD; n=25) vs. no LB pathology (n=53) with high sensitivity (100%) and specificity (94%). αSyn RT-QuIC thus correctly identified all clinicopathologically confirmed cases with standard LBD. It is worth noting that all 4 cases with DLB also met criteria for AD. In cases with atypical parkinsonian syndromes PSP (n=5) / CBS (n=1) / MSA (n=1), 6 out 7 were negative. The one αSyn RT-QuIC positive individual with an atypical parkinsonian syndrome had clinicopathological PSP with no evident LBs. It is also worth noting that the one case with neuropathologically confirmed MSA did not have any LB pathology and was αSyn RT-QuIC negative (Table 3).

**Table 3.** Distribution of neuropathological diagnosis in the (AZSAND/BBDP) cohort.

| Diagnostic category<br>NP diagnosis | N | αSyn<br>RT-QuIC<br>positive | αSyn<br>RT-QuIC<br>negative | Sensitivity | Specificity |
| --- | --- | --- | --- | --- | --- |
| **Standard LBD** | **25** | **25** |  | **100%** |  |
| DLB-AD | 4 | 4 |  | 100% |  |
| PD | 16 | 16 |  | 100% |  |
| PD-AD | 4 | 4 |  | 100% |  |
| PD-PSP* | 1 | 1 |  | 100% |  |
| **Non-standard LBD** | **23** | **13** | **10** | **57%** |  |
| ADLB | 14 | 9 | 5 | 64% |  |
| ILBD | 4 | 2 | 2 | 50% |  |
| LBS-PSP | 1 |  | 1 | 0% |  |
| LBS-VaD | 2 |  | 2 | 0% |  |
| LBS-VaD-PSP<br>** | 1 | 1 |  | 100% |  |
| LBS-Astrocytoma | 1 | 1 |  | 100% |  |
| **Lewy Body**<br>**negative** | **53** | **3** | **50** |  | **94%** |
| PSP | 2 | 1 | 1 |  | 50% |
| PSP-AD | 2 |  | 2 |  | 100% |
| VaD-CBD | 1 |  | 1 |  | 100% |
| MSA | 1 |  | 1 |  | 100% |
| AD | 7 | 2 | 5 |  | 71% |
| VaD-AD | 7 |  | 7 |  | 100% |
| Controls | 25 |  | 25 |  | 100% |
| Other | 8 |  | 8 |  | 100% |

\* included as PD in statistical analyses. \*\* Microscopic changes of PSP, not included as clinicopathological PSP in statistical analyses. Abbreviations: AD = Alzheimers disease; ADLB = Alzheimer's disease with Lewy bodies, not meeting criteria for DLB; CBD = Corticobasal degeneration; DLB = Dementia with Lewy Bodies; LBS = Lewy Bodies, LBD = Lewy Body Disorders; ILBD = Incidental Lewy Body Disease; VaD = Vascular dementia; MSA = Multiple System Atrophy; PSP = Supranuclear Palsy.

Of cases with “non-standard LBD” (i.e., AD with Lewy Bodies not meeting criteria for DLB or PD, and ILBD, n=23), only 57% cases were αSyn RT-QuIC positive (Table 3). αSyn RT-QuIC positive cases with non-standard LBD did not have significantly higher LB stage (p=0.174) or higher LB density (p=0.152) compared to αSyn RT-QuIC negative cases (Table 4). There was no significant difference in LB density in any of the 10 measured brain regions between αSyn RT-QuIC positive and negative cases with “non-standard LBD”. Further, there was no difference in UPDRS motor score or MMSE between αSyn RT-QuIC positive and negative cases with “non-standard LBD” (p=0.755, p=0.686 respectively).

**Table 4.** Lewy body stage by αSyn RT-QuIC status in Non-standard LBD.

|  | **αSyn RT-QuIC**<br>**positive** | **αSyn RT-QuIC**<br>**negative** |
| --- | --- | --- |
| **Stage I** | 1 | 2 |
| **Stage IIa** | 2 | 3 |
| **Stage IIb** | 7 | 4 |
| **Stage III** | 3 | 1 |
| **Stage IV** | 0 | 0 |

#### CSF αSyn RT-QuIC versus amyloid-β, tau and TDP-43 pathological changes, AZSAND/BBDP cohort

There were no significant differences in total amyloid-β plaque density score or total neurofibrillary tangle density score between αSyn RT-QuIC positive and negative cases. Further, there was no significant difference between the presence/absence of TDP-43 pathology between to αSyn RT-QuIC positive and negative cases (Table 2).

## Discussion

In this study we describe αSyn RT-QuIC testing of CSF from both a clinical cohort and a neuropathological cohort from a longitudinal clinicopathological study. Previous neuropathological studies with αSyn RT-QuIC of CSF have mainly investigated verified clinicopathological diagnoses with LBD vs. cases with no LBD, finding a high diagnostic accuracy in this setting [5, 14, 55], which we can confirm. In the present study we also investigated cases with “non-standard LBD”, i.e. those cases with neuropathological findings of LBD but not meeting criteria for PD or DLB. We found that CSF αSyn RT-QuIC reliably identified cases with LB stage III-IV (Fig. 1), cases with LB disease with cortical involvement (Fig. 2) and cases with high LB brain loads (Fig. 3) and separate these from cases with low LB density and no LBs with a high accuracy. However, the sensitivity is poor when investigating cases with modest LB pathology, defined either as i) low LB stage (I and IIa-IIb), or ii) limited spread of LBs restricted to the brainstem or amygdala, or iii) intermediate LB brain loads (56%, 50% and 64% respectively), but the number of cases were relatively small.

The diagnostic accuracy was very high for neuropathologically verified cases with standard LB disease vs. cases with no LBs at all with a sensitivity of 100% and a specificity of 94%, which is consistent with previous results [5, 14, 23, 55]. However, in parallel to the results with low diagnostic accuracy for cases with intermediate LB density or limited spread of LBs, the diagnostic accuracy was low in cases with “non-standard LBD”, with 57% being αSyn RT-QuIC positive. This is partially in line with the exploratory results by Fairfoul et al. who found up to 31% αSyn RT-QuIC positivity in AD cases with incidental LB, however the number with non-standard LBD in that study was very low [23]. In the present study, the αSyn RT-QuIC positive cases with “non-standard LBD” did not have significantly higher LB stage or LB density compared to the αSyn RT-QuIC negative cases with “non-standard LBD”. Further, there was no significant difference in LB density in any of the 10 different brain regions in αSyn RT-QuIC positive cases vs. αSyn RT-QuIC negative cases in the “non-standard LBD group”. It is therefore not possible to speculate, in this cohort, that the αSyn RT-QuIC positive cases with “non-standard LBD” would have been at higher risk of developing clinical Lewy body disease. However, there might be a threshold effect, where these cases are close to the threshold of αSyn RT-QuIC positivity/negativity, yielding a higher variability in the result. This is likely a result of lower total brain (and CSF) load of a-syn seeds in this group of cases with less widespread LB pathology seldom affecting the cortex. Further, the semiquantitative method of obtaining LB regional density score allows for some variability within a given density score. An alternative, but less likely and speculative, explanation is that the α-syn seeds present in the cortex are more prone to induce aggregation of monomeric α-syn.

The presence of more than one pathology is prevalent in neurodegenerative disorders [10, 36, 53] and Aβ and tau pathologies may act synergistically with α-syn pathology influencing the clinical presentation and prognosis in LBD [38]. Further, misfolded α-syn might potentiate aggregation of tau [6, 50]. However, the CSF αSyn RT-QuIC assay could in the present study specifically identify LB pathology without any associations to the load of Aβ, tau or TDP-43 pathology changes, although the number of cases investigated for TDP-43 pathology was low.

In our clinical cohort, we found a high sensitivity (95%) for αSyn RT-QuIC in LBD. However, the specificity in the present study was 83%, which is lower compared to previous results [28, 55]. One can speculate that the lower specificity in the clinical cohort reflects the fact that some of the individuals included as controls could have preclinical LBD. Indeed, 2 individuals initially included as controls that later converted to PD or DLB were both αSyn RT-QuIC positive. Further, the controls in the present study were mainly spouses or in some cases relatives of individuals with PD (mostly) or atypical parkinsonian disorders.

In our study, 33% of clinical MSA were αSyn RT-QuIC positive and 0/1 of the neuropathologically confirmed cases with verified MSA was αSyn RT-QuIC positive. This is in line with previous studies [55, 63]. It has been proposed that this is due to a different strain of α-syn in MSA [48, 49, 62], yielding a different kinetic profile [60], or no aggregation at all [55], in different αSyn seed amplification assays. Our assay was developed for the detection of seeds of Lewy body disorders rather than MSA, and others have shown better detection of MSA-associated seeds using alternative amplification conditions [42, 56]. Perhaps more surprising is our low specificity in PSP cases (67%) in the clinical cohort. However, clinical diagnoses are difficult and inherently uncertain, especially for atypical parkinsonian syndromes, and the presence of multiple pathologies are common in neurodegenerative disorders [36, 37, 53]. This makes the inclusion of a cohort of neuropathologically confirmed cases in the present study all the more important. In clinicopathological cases with PSP/CBD/MSA we found all but one case to be αSyn RT-QuIC negative; however, the number was small.

Overall, CSF αSyn RT-QuIC has become a promising biomarker assay for the diagnosis of PD and DLB that may vastly improve the diagnostic work up in these patients, but may not greatly improve the detection of individuals at the very early stages of the disease. Several studies have shown a high sensitivity for α-syn seed amplification assays in prodromal cases [35, 54, 55, 61]. Isolated rapid-eye-movement sleep behavior disorder (iRBD) is a common non-motor symptom in LBD and also a common prodromal symptom of LBD [12]. In a recent longitudinal study on patients with iRBD, 62% converted to PD or DLB during follow-up, of whom 97% were CSF αSyn RT-QuIC positive [35]. In the present study, αSyn RT-QuIC positivity was seen up to 5.5 years prior to diagnosis of the two LBD converters.

However, given the results in the present study with a very low diagnostic accuracy in cases with relatively low LB load (defined as either “non-standard LBD”, intermediate LB density or limited spread of LB pathology), further *longitudinal* studies are needed to clarify how early in the pre-symptomatic and prodromal phases individuals with early LBD convert to αSyn RT-QuIC positivity in CSF.

The possibility of accurate pre-symptomatic or prodromal diagnosis is especially relevant in coming trials on disease modifying therapies, as these are most likely to be effective if initiated early on in the disease course. αSyn RT-QuIC would likely be a useful test for at risk iRBD individuals in clinical drug trials to monitor the effect of disease modifying therapies initiated at the earliest stage.

A limitation to the present study is the lack of neuropathological data in the clinical cohort. However, we also studied αSyn RT-QuIC in a neuropathological cohort which strengthens the results. Further, participants in the clinical cohort are followed over time to ascertain as certain a diagnosis as possible. Indeed, the three αSyn RT-QuIC negative individuals clinical PD diagnosis were followed for 2-5 years in the study, showing typical features for PD including L-dopa response but were nonetheless early PD with a disease duration of ≤5 years.

In conclusion, the present study confirms that αSyn RT-QuIC has excellent sensitivity and specificity for cases with clinicopathologically verified LB disorders vs those with no LB pathological changes. However, the diagnostic accuracy was poor for cases with LB pathology restricted to non-cortical areas and having an overall low-to-intermediate LB brain load. Further studies are needed to investigate what factors influence αSyn RT-QuIC results in these cases with more limited LB pathological changes and determine at what timepoint in the pre-symptomatic or prodromal stages of LBD that the αSyn RT-QuIC result converts from sub-threshold to clear positivity.

## Data Availability

Anonymized data will be shared by request from a qualified academic investigator for the sole purpose of replicating procedures and results presented in the article and as long as data transfer is in agreement with EU legislation on the general data protection regulation and decisions by the Ethical Review Board of Sweden and Region Skane, which should be regulated in a material transfer agreement.

## Acknowledgements

The authors thank the study participants and their families. Without their invaluable contribution, this study would not have been possible.

Work at the authors’ research centres was supported by the Swedish Research Council (2016-00906), the Knut and Alice Wallenberg foundation (2017-0383), the Marianne and Marcus Wallenberg foundation (2015.0125), the Strategic Research Area MultiPark (Multidisciplinary Research in Parkinson’s disease) at Lund University, the Swedish Alzheimer Foundation (AF-939932), the Swedish Brain Foundation (FO2021-0293), The Parkinson foundation of Sweden (1280/20), the Konung Gustaf V:s och Drottning Victorias Frimurarestiftelse, the Skåne University Hospital Foundation (2020-O000028), Regionalt Forskningsstöd (2020-0314) and the Swedish federal government under the ALF agreement (2018-Projekt0279). This work was supported in part by the Intramural Research Program of the NIAID, NIH. We are grateful to the Banner Sun Health Research Institute Brain and Body Donation Program of Sun City, Arizona for the provision of human biological materials. The Brain and Body Donation Program has been supported by the National Institute of Neurological Disorders and Stroke (U24 NS072026 National Brain and Tissue Resource for Parkinson’s Disease and Related Disorders), the National Institute on Aging (P30 AG19610 Arizona Alzheimer’s Disease Core Center), the Arizona Department of Health Services (contract 211002, Arizona Alzheimer’s Research Center), the Arizona Biomedical Research Commission (contracts 4001, 0011, 05-901 and 1001 to the Arizona Parkinson’s Disease Consortium) and the Michael J. Fox Foundation for Parkinson’s Research.

The funding sources had no role in the design and conduct of the study; in the collection, analysis, interpretation of the data; or in the preparation, review, or approval of the manuscript.

## Author contribution

Sara Hall: Execution of the research project. Design and execution of the statistical analysis. Writing the manuscript draft.

Christina Orrù: Execution of the research project and performing the αSyn RT-QuIC analyses. Co-writing the manuscript. Review and critique of statistical analysis.

Geidy E. Serrano: Execution of the research project. Review and critique of the statistical analysis. Review and critique of the manuscript.

Douglas Galasko: Organization of the research project. Review and critique of statistical analysis and manuscript.

Andrew G. Hughson: Execution of the research project. Review and critique of statistical analysis and manuscript.

Bradley R. Groveman: Execution of the research project. Review and critique of statistical analysis and manuscript.

Charles H. Adler: Organization of the research project. Review and critique of statistical analysis and manuscript.

Thomas G. Beach: Organization of the research project. Provision of biospecimens. Review and critique of statistical analysis and manuscript.

Byron Caughey: Conception and organization of the research project. Review and critique of the manuscript.

Oskar Hansson: Initiation and design of the study. Conception and organization of the research project. Design and review and critique of statistical analysis. Review and critique of the manuscript.

